# Cohort profile: the Swiss Cerebral Palsy Registry (Swiss-CP-Reg) cohort study

**DOI:** 10.1101/2021.11.02.21265824

**Authors:** Fabiën N. Belle, Sandra Hunziker, Joël Fluss, Sebastian Grunt, Stephanie Juenemann, Christoph Kuenzle, Andreas Meyer-Heim, Christopher J. Newman, Gian Paolo Ramelli, Peter Weber, Claudia E. Kuehni, Anne Tscherter

**Affiliations:** Institute of Social and Preventive Medicine, University of Bern, Bern, Switzerland; Center for Primary Care and Public Health (Unisanté), University of Lausanne, Lausanne, Switzerland; Paediatric Neurology Unit, University Children’s Hospital Geneva, Geneva, Switzerland; Division of Neuropediatrics, Development and Rehabilitation, Department of Pediatrics, Inselspital Bern, University Hospital, University of Bern, Switzerland; Division of Neuropaediatric and Developmental Medicine, University Children’s Hospital of Basel (UKBB), University of Basel, Basel, Switzerland; Children’s Hospital of Eastern Switzerland, St Gallen, Switzerland; Swiss Children’s Rehab, University Children’s Hospital Zurich, Affoltern am Albis, Switzerland; Paediatric Neurology and Neurorehabilitation Unit, Lausanne University Hospital (CHUV), Lausanne, Switzerland; University of Lausanne (UNIL), Lausanne, Switzerland; Gian Paolo Ramelli, Neuropaediatric Unit, Paediatric Institute of Southern Switzerland, Ospedale San Giovanni, Bellinzona, Switzerland; Children’s University Hospital, Inselspital, University of Bern, Bern, Switzerland

**Keywords:** Cerebral Palsy, Swiss Cerebral Palsy Registry, Patient registry, Paediatrics, Epidemiology, Europe

## Abstract

**BACKGROUND:** Cerebral Palsy (CP) is a group of permanent disorders of movement and posture that follows injuries to the developing brain. It results in motor dysfunction and a wide variety of comorbidities like epilepsy, pain, speech, hearing and vision disorders, cognitive dysfunction, and eating and digestive difficulties. Central data collection is essential to study the epidemiology, clinical presentations, care, and quality of life of patients affected by CP. CP specialists founded the Swiss Cerebral Palsy Registry (Swiss-CP-Reg) in 2017. This paper describes the design, structure, aims and achievements of the Swiss-CP-Reg and presents first results.

**METHODS:** Swiss-CP-Reg records patients of any age suffering from CP who are born, treated, or live in Switzerland. It collects data from medical records and reports, from questionnaires answered by patients and their families, and from data linkage with routine statistics and other registries. The registry contains information on diagnosis, clinical presentation, comorbidities, therapies, personal information, family history, and quality of life.

**Results:** From August 2017 to August 2021, 546 participants (55% males, mean age at registration 8 years (interquartile range [IQR]: 5-12) were enrolled in Swiss-CP-Reg. Most had been born at term (56%), were less than 2 years old at diagnosis (73%, median 9 months, IQR: 18-25), and were diagnosed with spastic CP (76%). Most (59%) live with a mild motor impairment (Gross Motor Function Classification System [GMFCS] level I or II), 12% with a moderate motor impairment (GMFCS level III), and 29% with a severe motor impairment (GMFCS level IV or V). In a subset of 170 participants, we measured intelligence quotient (IQ) and saw a reduced IQ by GMFCS scale increase. The Swiss-CP-Reg has a strong interest in research with currently 5 nested projects running, and many more planned.

**CONCLUSIONS:** Swiss-CP-Reg collects and exchanges national data on people living with CP to answer clinically relevant questions. Its structure enables retrospective and prospective data collection and knowledge exchange between experts to optimise and standardise treatment and improve the health and quality of life of those diagnosed with CP in Switzerland.

**ClinicalTrials.gov identifier:** NCT04992871

## INTRODUCTION

Cerebral palsy (CP) is a group of permanent disorders of movement, posture, and motor function causing activity limitation. With a prevalence of 1.5 to 2.5 per 1’000 live-born children [1], it is the most common cause of physical disability in children. It is estimated that approximately 3000 children and adolescents and 12’000 adults live in Switzerland with CP based on international data and extrapolated data from the eastern part of the country (St. Gallen) [2]. CP results from a non-progressive lesion or brain malformation that occurs during the prenatal, perinatal, or postnatal period, e.g., ischemic lesions of the neonatal brain or genetic predispositions leading to brain malformation [3]. CP shows a large variability in type and distribution of movement abnormality and degree of functional impairment. Besides motor dysfunction, persons with CP suffer from many comorbidities, including epilepsy, speech, hearing or vision disorders, cognitive dysfunction, eating and digestive difficulties, behavioural disorders, and secondary musculoskeletal problems including associated pain [1,3-5]. These comorbidities can restrict persons with CP in various dimensions of social life, such as education, play and sport activities, and work participation [6-10]. Within Switzerland, only 20% of adults with CP have paid employment in the primary labour market job [11]. Besides these restrictions persons with CP experience also their family’s everyday life is influenced by their disability. The CP of a child can impact the (mental) health and well-being of affected parents [12-18] and limits siblings in their activities [19,20]. Currently, there is no cure for CP and patients often need lifelong support by an interdisciplinary team of specialists.

The involvement of an interdisciplinary team including many different specialists makes it difficult for researchers to obtain the full spectrum of patient data. A centralised data collection, where all information on people with CP can be linked, is therefore essential to optimize and standardise care, improve clinical surveillance, develop preventive strategies for comorbidities, and study the quality of life of affected people. Other countries have built up regional [21-23], or national [24-31] registries, or participate in networks [32,33]. In Switzerland, knowledge about CP in general and knowledge of supportive treatment in particular, is sparse. Previous research on CP in Switzerland included regional projects that focused on specific questions, such as epidemiologic research, work on neuroimaging and health related quality of live in unilateral spastic CP after neonatal arterial ischemic stroke [34-36], interventions to improve upper limb function in unilateral spastic CP [37-39], the early diagnosis of cerebral palsy in infants at risk [40], robot-assisted rehabilitation [41-43], hip reconstruction [44], gait analysis in children and young adult with CP [45,46], application patterns of physiotherapy (continuous vs. blocks) [47], and behavioural and emotional problems [48]. Data has also been contributed to international studies [40,41,49,50]. In 2017 CP specialists from the entire country decided on a national collaboration and founded the Swiss Cerebral Palsy Registry (Swiss-CP-Reg) to collect national data and promote CP research in Switzerland. Swiss-CP-Reg aims to optimise treatment and improve the health and quality of life of children living with CP by establishing a platform for research and knowledge exchange between patients, healthcare providers, researchers, and other stakeholders. This paper describes the design, structure, aims, and achievements of the Swiss-CP-Reg and presents baseline characteristics of participants.

## MATERIALS AND METHODS

### Study design

Swiss-CP-Reg is a national patient registry (ClinicalTrials.gov registration number: NCT04992871) that collects medical and socio-demographic data on people with CP. Persons living with CP who give their consent are registered in Swiss-CP-Reg, and their data is currently collected at regular intervals at the paediatric clinics in Basel, Bellinzona, Bern, Geneva, Lausanne, St. Gallen, and Zurich. In the coming years, it will extend data collection to include smaller hospitals and medical practices and invite adults to participate.

#### Aims of the Swiss-CP-Reg

The aims of the registry are, to

1. Identify all children, adolescents and adults living with CP across the country, characterize their phenotypes and determine incidence, prevalence, time trends and regional differences.
2. Document diagnostic evaluations, treatments, quality of life, morbidity, mortality, and risk factors.
3. Establish a research platform for clinical, epidemiological, and basic research to support participant recruitment for population based as well as interventional studies and to answer questions on topics like health, health care, education, social aspects, and quality of life.
4. Establish a platform for knowledge exchange between clinics, researchers, therapists, national and cantonal health authorities, and international parties.

### Study population and recruitment procedure

Swiss-CP-Reg registers all children, adolescents, and adults diagnosed with CP who have been born, are treated, or live in Switzerland. The diagnosis is made based on the Surveillance of Cerebral Palsy in Europe (SCPE) decision tree [32]. To assure that only children who meet the inclusion criteria are included, registration of participants is restricted to paediatricians specialised in neurology, rehabilitation, or development and a confirmation of the diagnosis is required at the age of 5 years in case children are enrolled at an earlier age. We exclude participants with pure muscular hypotonia, neurometabolic diseases (e.g., neuronal storage diseases, leukodystrophies) and other progressive neurological diseases (e.g., spinocerebellar ataxias, hereditary spastic paraplegia, Rett syndrome, epileptic encephalopathy).

Physicians in clinics or practices identify eligible participants according to their clinical diagnosis and recruit them during routine medical consultations (**Figure 1**). Physicians inform eligible participants and caregivers and collect informed consent during their consultations. Study information and consent forms are available in four languages, French, German, Italian, and English. Those who provide informed consent are registered in the Swiss-CP-Reg. For those who do not consent, the registry collects an anonymized minimal data set consisting of year of birth, year of death (if applicable), sex, gestational age, birth weight, and CP type (**Table 1**).

**Figure 1.**
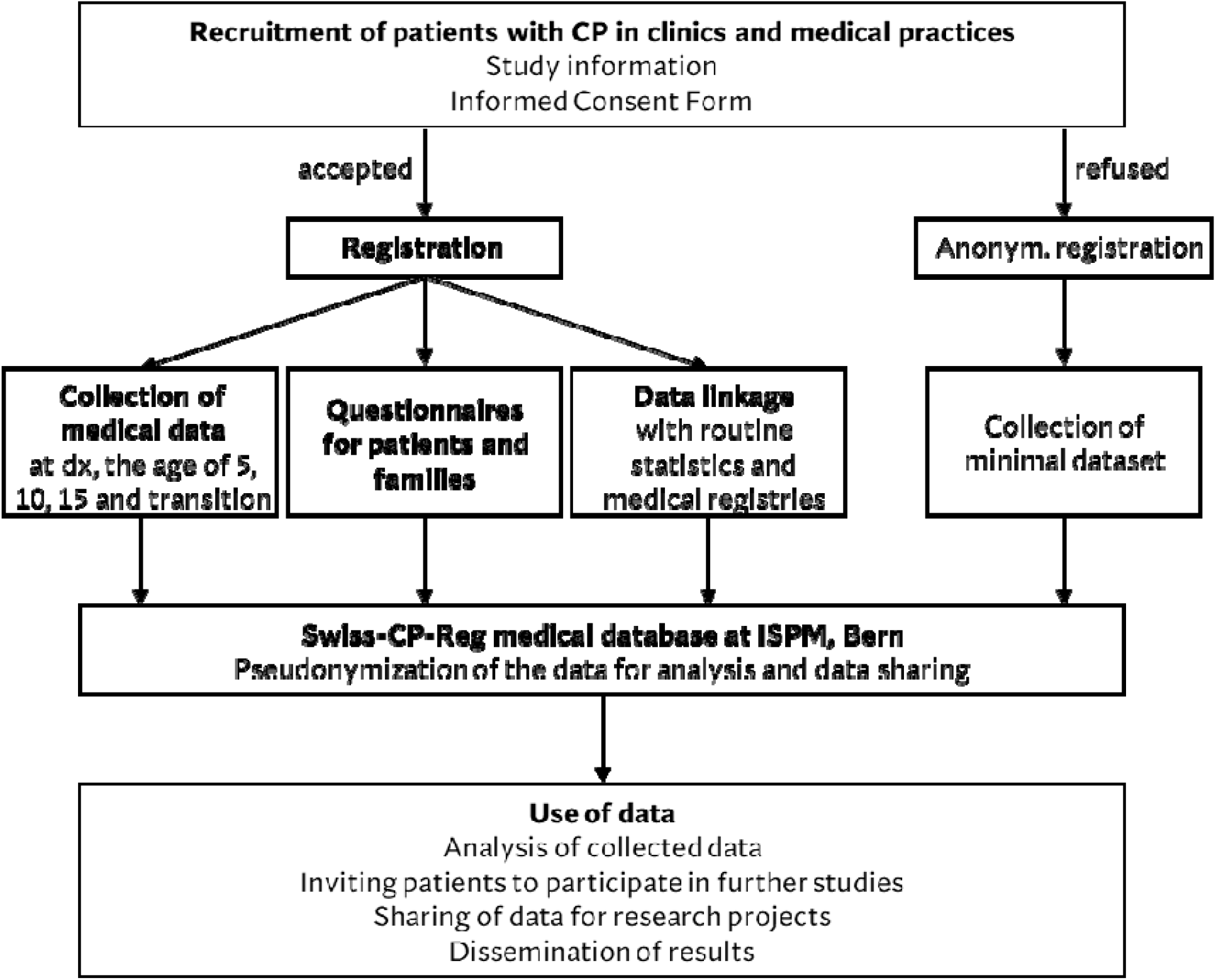
Schematic chart of patient recruitment, data collection, and use of data of the Swiss Cerebral Palsy Registry. CP, cerebral palsy; dx, diagnosis; ISPM, institute of social and preventive medicine; transition, transition to adult care.

**Table 1.**
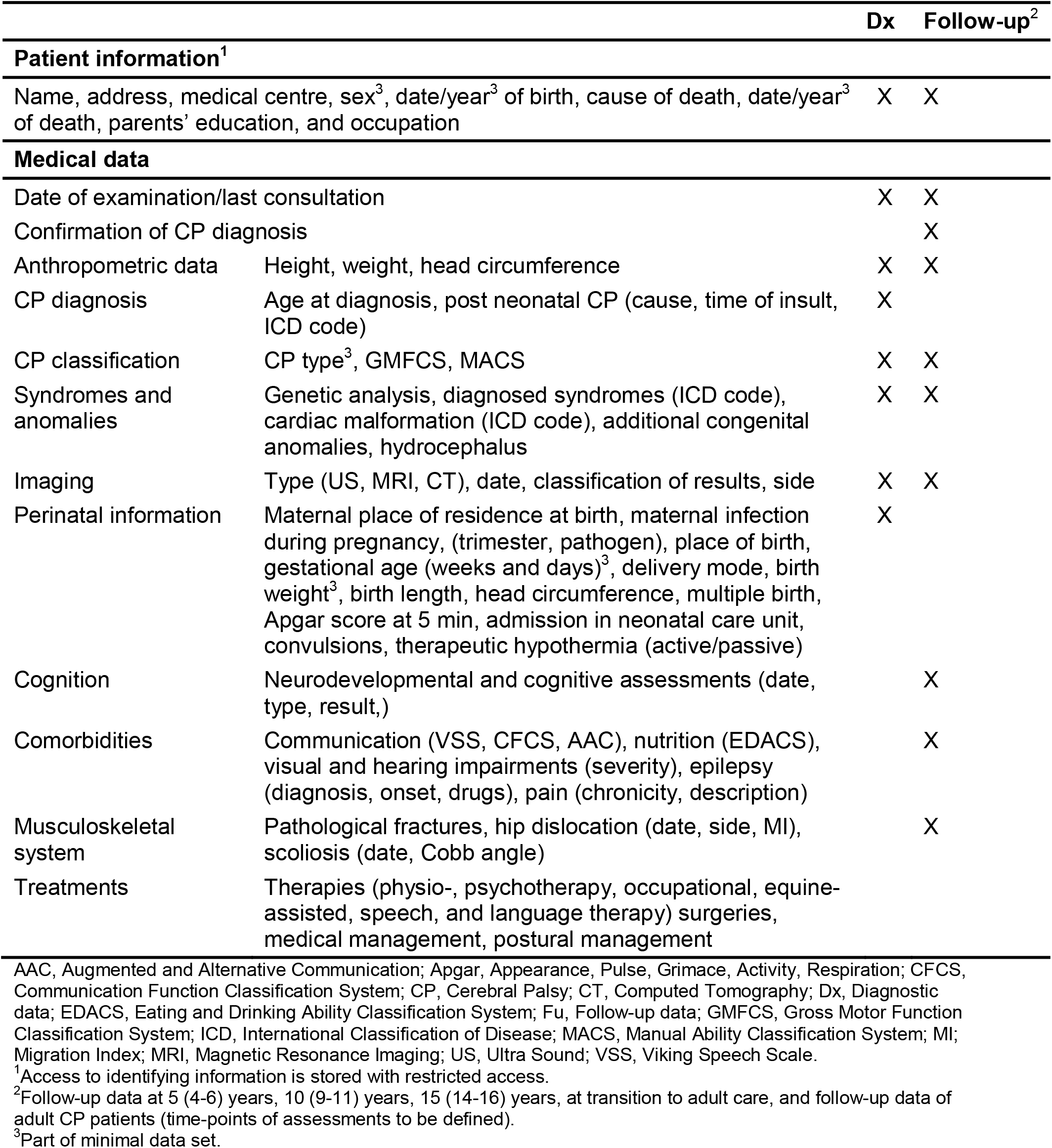
Description of the data collected in the Swiss Cerebral Palsy Registry and time points of assessment

|  |  | Dx | Follow-up<sup>2</sup> |
| --- | --- | --- | --- |
| **Patient information<sup>1</sup>** |  |  |  |
| Name, address, medical centre, sex<sup>3</sup>, date/year<sup>3</sup> of birth, cause of death, date/year<sup>3</sup> of death, parents' education, and occupation |  | X | X |
| **Medical data** |  |  |  |
| Date of examination/last consultation |  | X | X |
| Confirmation of CP diagnosis |  |  | X |
| Anthropometric data | Height, weight, head circumference | X | X |
| CP diagnosis | Age at diagnosis, post neonatal CP (cause, time of insult, ICD code) | X |  |
| CP classification | CP type<sup>3</sup>, GMFCS, MACS | X | X |
| Syndromes and anomalies | Genetic analysis, diagnosed syndromes (ICD code), cardiac malformation (ICD code), additional congenital anomalies, hydrocephalus | X | X |
| Imaging | Type (US, MRI, CT), date, classification of results, side | X | X |
| Perinatal information | Maternal place of residence at birth, maternal infection during pregnancy, (trimester, pathogen), place of birth, gestational age (weeks and days)<sup>3</sup>, delivery mode, birth weight<sup>3</sup>, birth length, head circumference, multiple birth, Apgar score at 5 min, admission in neonatal care unit, convulsions, therapeutic hypothermia (active/passive) | X |  |
| Cognition | Neurodevelopmental and cognitive assessments (date, type, result,) |  | X |
| Comorbidities | Communication (VSS, CFCS, AAC), nutrition (EDACS), visual and hearing impairments (severity), epilepsy (diagnosis, onset, drugs), pain (chronicity, description) |  | X |
| Musculoskeletal system | Pathological fractures, hip dislocation (date, side, MI), scoliosis (date, Cobb angle) |  | X |
| Treatments | Therapies (physio-, psychotherapy, occupational, equine-assisted, speech, and language therapy) surgeries, medical management, postural management |  |  |

AAC, Augmented and Alternative Communication; Apgar, Appearance, Pulse, Grimace, Activity, Respiration; CFCS, Communication Function Classification System; CP, Cerebral Palsy; CT, Computed Tomography; Dx, Diagnostic data; EDACS, Eating and Drinking Ability Classification System; Fu, Follow-up data; GMFCS, Gross Motor Function Classification System; ICD, International Classification of Disease; MACS, Manual Ability Classification System; MI, Migration Index; MRI, Magnetic Resonance Imaging; US, Ultra Sound; VSS, Viking Speech Scale.
<sup>1</sup>Access to identifying information is stored with restricted access.
<sup>2</sup>Follow-up data at 5 (4-6) years, 10 (9-11) years, 15 (14-16) years, at transition to adult care, and follow-up data of adult CP patients (time-points of assessments to be defined).
<sup>3</sup>Part of minimal data set.

### Data collection and data sources

The Swiss-CP-Reg collects data through three main paths: 1) medical records from clinics and private practices; 2) questionnaires sent to patients and families; and 3) re-use of data from routine statistics and other registries.

#### Medical data

Baseline medical data is collected in the participating clinics with the support of the registry team at the ISPM in Bern (**Figure 1, Figure 2**). The variable list is based on the SCPE dataset [51] and includes additional information on specific topics such as therapies, scoliosis, hip surveillance and pain (**Table 1**). Personal information collected from the medical records includes patient’s name, contact information, date of birth, and contact information of the treating paediatrician. This is necessary to assure long term follow-up of the patients over decades and to invite patients and families to surveys and clinical studies. The dataset enables a detailed description of people living with CP, their clinical condition, diagnosis, measures of severity, and care. CP is diagnosed according to the SCPE guidelines, and severity and comorbidities are assessed using standardized tools such as Gross Motor Function Classification System (GMFCS), Manual Ability Classification System (MACS), or Communication Function Classification System (CFCS). The use of these instruments allows to record the change of the condition over time and to compare centres. Medical data are collected at regular intervals (**Figure 2, Table 1**). The main time-periods when data are collected are: at diagnosis (independent of age), at the age of 5, 10, and 15 years, and at the time of transition to adult care. Follow-up during adulthood is foreseen. When a patient is included in the Swiss-CP-Reg, available data up to the time point of enrolment is collected retrospectively. From then onwards, follow-up data is collected prospectively until death or loss to follow-up. Because of limited resources, centres now collect all available medical data from birth onwards for young children born after 2009. For adolescents born before 2010, we currently collect CP classification and severity data (GMFCS and intelligence quotient [IQ]). A retrospective data collection from birth until enrolment will follow for adolescents as soon as resources allow.

**Figure 2.**
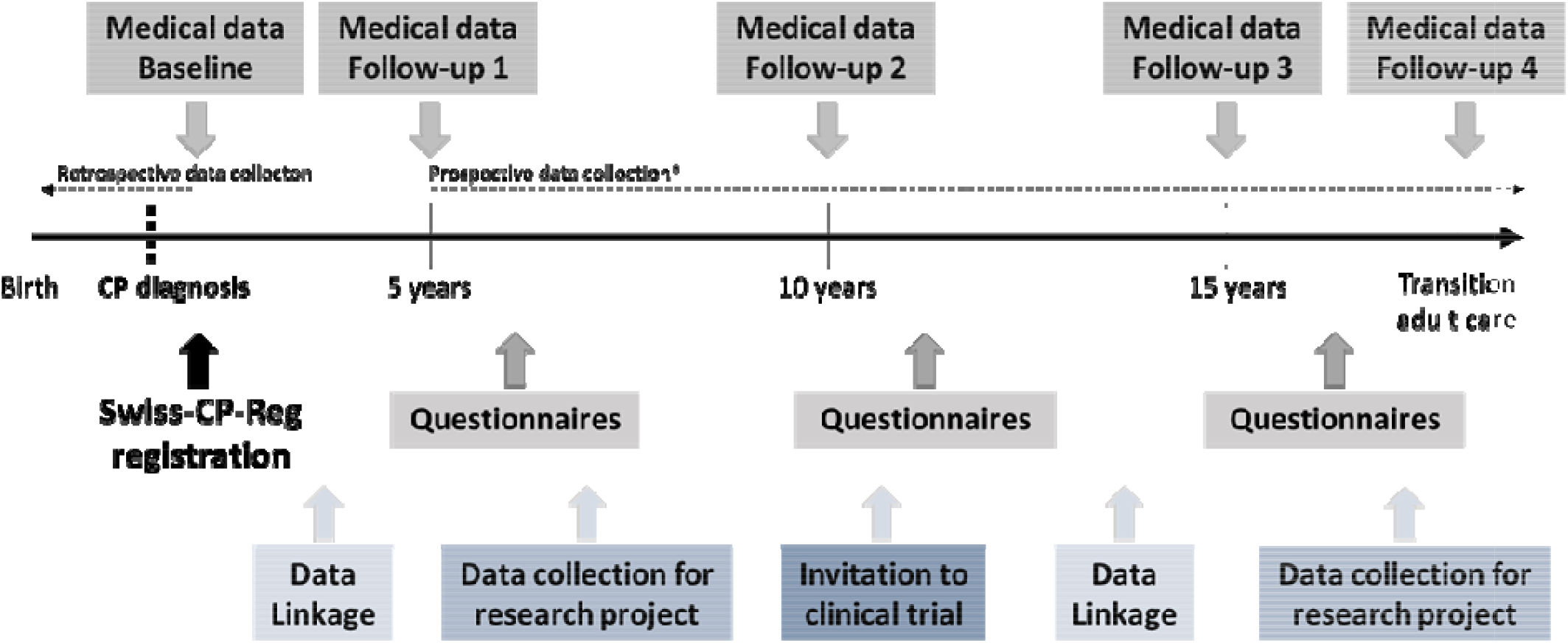
Scematic flow chart of Swiss-CP-Reg; data collection management and timelines. CP, cerebral palsy; Swiss-CP-Reg, Swiss Cerebral Palsy Registry * Medical follow-up data can be collected retrospectively if a participant is registered after diagnosis

#### Questionnaire data

Questionnaires are sent to people with CP in the registry and their families at regular intervals. Participation is voluntary and can be accepted or declined for each survey. Questionnaires cover different topics including healthcare, nutrition, sleep, pain, use of medical equipment, use of ancillary services, academic information including early childhood education level, school and professional integration, family history, health behaviour, quality of life, daily life participation, and needs and concerns of patients and families.

#### Re-use of data from routine statistics and other registries

data from routine statistics and other registries can be linked to the Swiss-CP-Reg to answer specific research questions. For example, the Federal Statistics Office (FSO) Live Birth Registry can provide information on gestational age, birth weight, birth length, and parental age; the FSO Cause of Death and Stillbirth Statistics can contribute causes of death; Hospital Episode Statistics can inform on length of hospital stay; the Swiss Neonatal Network and Follow-up Group (SwissNeoNet) has information on the perinatal period for neonatal care data; and the Swiss Neuropediatric Stroke Registry includes infarct data. Authorisation for data linkage is needed from the data providers, e.g., from FSO or SwissNeoNet, for each analysis.

### Database management and data flows

The Swiss-CP-Reg database is built and managed using Research Electronic Data Capture (REDCap; Nashville, TN, USA) hosted at ISPM [52,53]. REDCap is a secure, web-based software platform supporting data capture for research studies. Captured personal data are strictly confidential. Access to the ISPM server and the Swiss-CP-Reg database is only given to authorised personnel and data are handled with uttermost discretion. Daily, weekly, and monthly back-ups of the database are performed and securely stored on the ISPM servers.

Most data are entered in the REDCap database directly by the local healthcare professionals or by members of the Swiss-CP-Reg team (**Figure 2**). The Swiss-CP-Reg team enters additional information that is sent to the ISPM via post or a secured e-mail address. Paper forms are securely locked at ISPM and can only be accessed by Swiss-CP-Reg team members. All paper forms, except for filled-out questionnaires, consent forms, and withdrawals, will be destroyed after digitalization and storage in the REDCap database.

### Data quality, analyses, and data sharing

Patient’s data is checked manually by the Swiss-CP-Reg team and by REDCap for completeness, plausibility, and consistency. Ambiguities are resolved in collaboration with treating healthcare professionals at the time of data entry or after the manual quality check. We randomly perform control checks to detect systematic mistakes during data entry. In addition, selected data points are cross-checked for plausibility with previously entered data. Data analyses are performed using dedicated programs (STATA, R).

Pseudonymised data can be made available to other research projects when legal requirements are met. This includes data usage by regional, national, or international research projects. Researchers interested in collaborative work can contact the Swiss-CP-Reg team to discuss planned projects or analyse existing data. The decision on collaboration is made by the Steering Board of the Swiss-CP-Reg.

### Governance, organisational structure, and funding

#### Organisational structure

In 2014 the setup of a national registry for CP in Switzerland was initiated by Dr. med. Christoph Kuenzle (Ostschweizer Kinderspital, St. Gallen). The setup was divided into three phases. In the preparation phase we defined the stakeholders, organizational structure, fundraising strategy, objectives, dataset, in- and exclusion criteria, participant recruitment, and data collection. During the following build-up phase, we applied for ethical approval, developed data transfer and use agreements and data collection guidelines, set up a REDCap database and dissemination tools, and requested funding. During the third phase, we started to recruit participants and collect data. We developed data quality checks and revised the initial dataset where needed.

We developed an inclusive and broad organizational structure, which comprises several specialised bodies. The *steering board* includes specialised paediatricians from all seven large Swiss child hospitals. The involvement of the steering board led to a harmonised national data collection and a more complete recording of a child’s clinical picture. The *operational management* is located at ISPM Bern and supports the data providers in recruitment and data collection. It hosts and maintains the database, takes care of the legal aspects and public relations, and promotes research. The *expert groups* include specialists from different fields, who support the registry, such as geneticists, paediatric orthopaedic surgeons, physiotherapists, speech therapists, nutritionists, and parents of children or adolescents with CP. The *general assembly* includes representatives of clinical centres, national patient organisations, medical societies, and expert groups. They meet once per year to discuss ongoing research and promote study participation. The *clinical centres* provide support in data collection.

Swiss-CP-Reg collaborates closely with the Swiss Research Platform for Paediatric Registries (SwissPedReg). This platform provides advice to registries in a build-up phase on for example the ethics application process, datasets and information collection, and organisational structure. Swiss-CP-Reg collaborates closely with national stakeholders, including patients and their families, patient organisations, clinics, medical societies, experts from various disciplines and regions, researchers, and international networks. All national stakeholders are included in the organisational structure of the registry. Swiss-CP-Reg also exchanges information with the collaborative network of CP registries across Europe (SCPE) and will apply for membership as soon as it reaches the prerequisites set by the SCPE e.g., the recruitment of patients also in small clinics and private practices.

#### Funding

The Swiss-CP-Reg (salaries, consumables, equipment) is financed by several funding bodies that include the ‘Schweizerische Stiftung für das cerebral gelähmte Kind (Stiftung Cerebral)’, Anna Mueller Grocholski Foundation, Swiss Academy of Childhood Disability (SACD), ‘Hand in Hand Anstalt’, ‘Ostschweizer Kinderspital’, the ACCENTUS Charitable Foundation (Walter Muggli Fund), and the Children’s Research Centre (University Children’s Hospital Zurich), Stiftung Cerebral was the main sponsor during the build-up phase of the registry.

### Ethical approval

In 2017, Swiss-CP-Reg obtained authorisation from the Cantonal Ethics Committee of Bern (2017-00873, risk category A, observational study) to collect national medical data from hospitals, clinics, and private practices, self-reported data from patients and families, to link data, and to retrospectively register deceased patients and patients lost to follow-up from clinics.

## RESULTS

### Status of patient registration and data collection

Active patient enrolment started in August 2017 in St. Gallen, followed in 2018 by Basel, Bellinzona, and Bern, in 2019 by Zurich and Geneva, and in 2020 by Lausanne. The Covid-19 pandemic slowed down recruitment in 2020 and 2021. By end of August 2021, Swiss-CP-Reg included data from 515 children and adolescents (0-17 years) and 31 adults (≥18 years), which represents an estimated 17% of all eligible children with CP in Switzerland. Among eligible participants who were identified and approached, only 30 (5%) refused participation and 4 (1%) were non-responders. For them we collected an anonymized minimal data set (**Table 2**). For about 80 patients the informed consent is pending; therefore, these patients have not yet been registered. By August 2021, we have collected baseline data at time of diagnosis for 400 participants, 5-year follow-up data for 266, 10-year follow-up data for 162, 15-year follow-up data for 31, and follow-up data at the timepoint of transition to adult care for two participants.

**Table 2.**
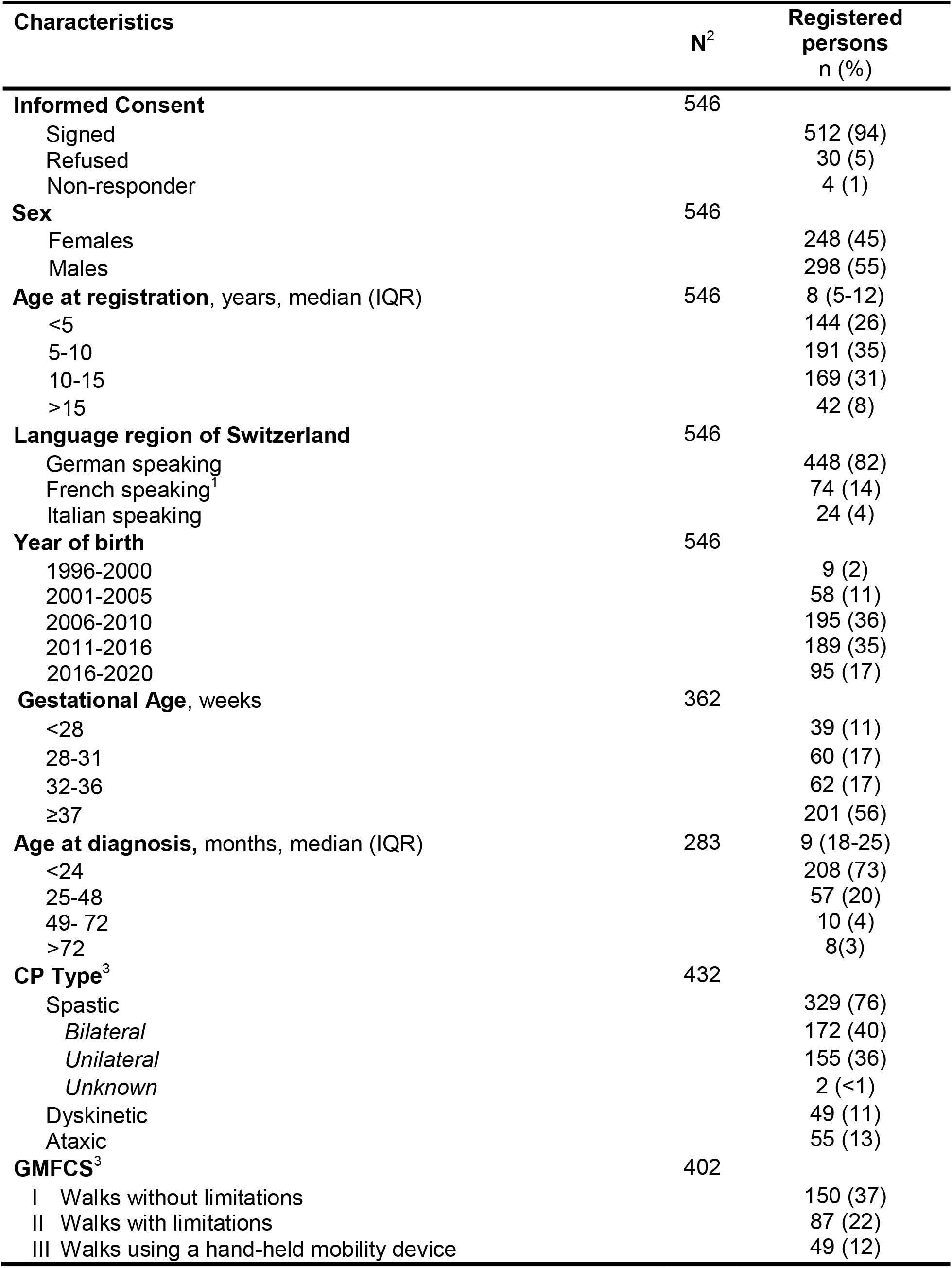
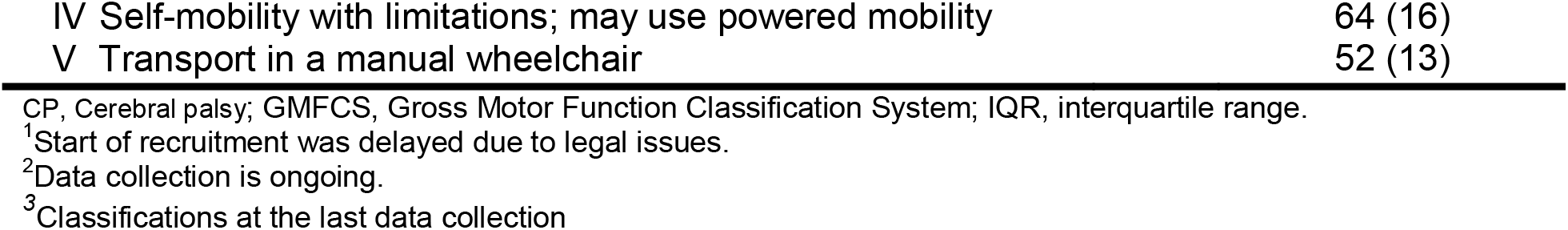
Characteristics of study participants in the Swiss Cerebral Palsy Registry (N=546, status at 31.08.2021)

| Characteristics | N<sup>2</sup> | Registered persons<br>n (%) |
| --- | --- | --- |
| **Informed Consent** | 546 |  |
| Signed |  | 512 (94) |
| Refused |  | 30 (5) |
| Non-responder |  | 4 (1) |
| **Sex** | 546 |  |
| Females |  | 248 (45) |
| Males |  | 298 (55) |
| **Age at registration, years, median (IQR)** | 546 | 8 (5-12) |
| <5 |  | 144 (26) |
| 5-10 |  | 191 (35) |
| 10-15 |  | 169 (31) |
| >15 |  | 42 (8) |
| **Language region of Switzerland** | 546 |  |
| German speaking |  | 448 (82) |
| French speaking<sup>1</sup> |  | 74 (14) |
| Italian speaking |  | 24 (4) |
| **Year of birth** | 546 |  |
| 1996-2000 |  | 9 (2) |
| 2001-2005 |  | 58 (11) |
| 2006-2010 |  | 195 (36) |
| 2011-2016 |  | 189 (35) |
| 2016-2020 |  | 95 (17) |
| **Gestational Age, weeks** | 362 |  |
| <28 |  | 39 (11) |
| 28-31 |  | 60 (17) |
| 32-36 |  | 62 (17) |
| ≥37 |  | 201 (56) |
| **Age at diagnosis, months, median (IQR)** | 283 | 9 (18-25) |
| <24 |  | 208 (73) |
| 25-48 |  | 57 (20) |
| 49- 72 |  | 10 (4) |
| >72 |  | 8(3) |
| **CP Type<sup>3</sup>** | 432 |  |
| Spastic |  | 329 (76) |
| *Bilateral* |  | 172 (40) |
| *Unilateral* |  | 155 (36) |
| *Unknown* |  | 2 (<1) |
| Dyskinetic |  | 49 (11) |
| Ataxic |  | 55 (13) |
| **GMFCS<sup>3</sup>** | 402 |  |
| I Walks without limitations |  | 150 (37) |
| II Walks with limitations |  | 87 (22) |
| III Walks using a hand-held mobility device |  | 49 (12) |

|  |  |
| --- | --- |
| IV Self-mobility with limitations; may use powered mobility | 64 (16) |
| V Transport in a manual wheelchair | 52 (13) |

CP, Cerebral palsy; GMFCS, Gross Motor Function Classification System; IQR, interquartile range.
<sup>1</sup>Start of recruitment was delayed due to legal issues.
<sup>2</sup>Data collection is ongoing.
<sup>3</sup>Classifications at the last data collection

### Characteristics of the study population

Table 2 summarises the basic socio-demographic and medical characteristics of the 546 registered participants. Slightly more than half were males (55%) with a median age at registration of 8 years (interquartile range [IQR]: 5-12 years). Most lived in the German speaking language region (82%); were born during the last ten years (year of birth 2011-2020, 52%), at term (gestational age ≥37 weeks, 56%), and were younger than 2 years at diagnosis (73%, median age 9 months, IQR: 18-25). The most prevalent type of diagnosis was spastic CP (76%) of whom 40% had bilateral and 36% had unilateral CP. Most (59%) live with a mild motor impairment (Gross Motor Function Classification System [GMFCS] level I, walks without limitations or II, walks with limitations). A moderate motor impairment was diagnosed in 12% (GMFCS level III, walks using a hand-held mobility device), and 29% had a severe motor impairment (GMFCS level IV, self-mobility with limitations; may use powered mobility or V, transport in a manual wheelchair). In a subset of 170 children and adolescents we had collected information on the intelligence quotient (IQ). There was a clear decrease of IQ with increasing level of GMFCS (**Figure 3**).

**Figure 3.**
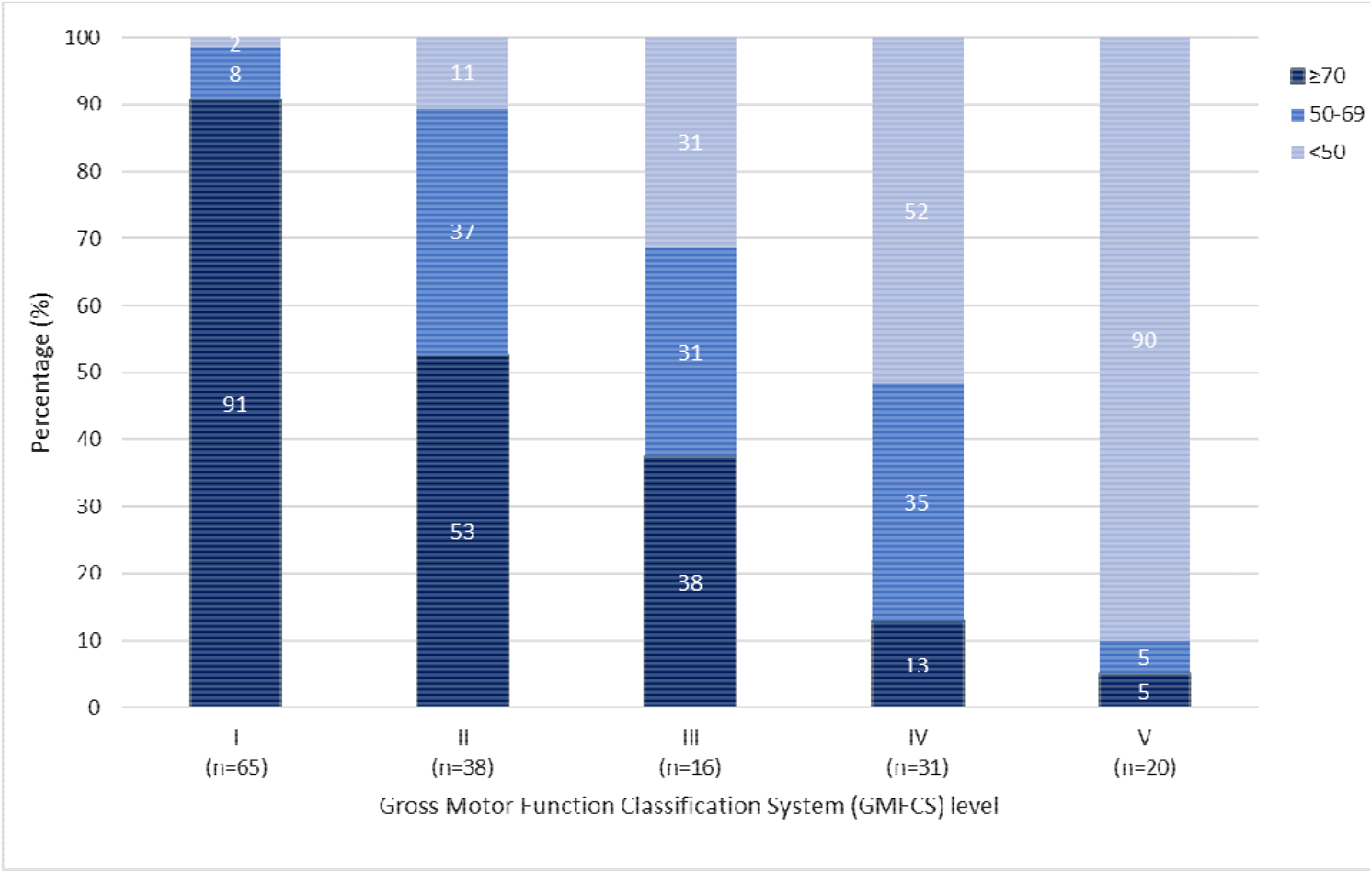
Intelligence quotient distribution, by Gross Motor Function Classification System, in a subset of 170 child and adolescent included in the Swiss-CP-Reg. Gross Motor Function Classification System (GMFCS) is subdivided in level I, Walks without limitations; level II, Walks with limitations; level III, Walks using a hand-held mobility device; level IV, Self-Mobility with limitations; may use powered mobility, and level V, Transported in a manual wheelchair; intelligence quotient (IQ). The P-value calculated from Chi-Square statistics comparing GMFCS levels (2-sided) is <0.001.

### Research projects

The Swiss-CP-Reg collaborates with experts from various fields to realize research projects that aim to standardize and improve CP therapies. It supports the monitoring and research of a wide range of outcomes and data exchange via linkages with national and regional projects led by interested investigators. Ongoing projects address the following broad range of topics: medical cannabinoids, needs and wishes of patients and families, hip surveillance in a variety of Swiss paediatric clinics, risk factors and outcomes of unilateral CP and data linkage with SwissNeoNet (**Table 3**). In addition, master students conduct projects within the scope of Swiss-CP-Reg. Three master theses have been carried out addressing nutrition, the epidemiology of epilepsy, and magnetic resonance imaging classification, and two are currently addressing the relation between neuroimaging patterns and upper limb function in unilateral spastic CP and the timing, type and treatment of epileptic seizures and pain via a survey (SG, CK) Liliane Raess evaluated in her doctoral thesis (under supervision of AMH and Prof. Dr. Hubertus van Hedel) the perinatal history, imaging, outcome and treatment of the Zurich cohort of children with CP and compared this cohort to other registries. Second aim was to investigate the differences in early history and diagnostic process between the rare and common neurologic subtypes of CP in the Zurich cohort.

**Table 3:** Ongoing and finalised research projects and collaborations of Swiss Cerebral Palsy Registry (September 2021)

| Principal investigator, place | Project |
| --- | --- |
| PD Dr. med. S. Grunt, Bern | **Prescription practices of medical cannabinoids in children with CP:** Medical cannabinoids are prescribed to children with CP despite limited evidence. The aim of the study is to assess the prescription practices of cannabinoids in children with CP with a particular focus on indications and preparations used and how well cannabinoids are tolerated. Data are collected from physicians by an online survey. |
| PD Dr. A. Tscherter, Bern | **Needs and wishes of patients with CP and their families:** Data on needs and wishes of children with CP and their families living in Switzerland is collected via questionnaires. The aim of this study is to uncover sources of concern by patients and families (such as quality of life, nutrition, communication, use of aids). |
| Prof. Dr. med. T. Dreher, Zurich | **Hip surveillance:** Monitoring the hips of patients with CP is important to detect and treat luxation at an early stage. Several countries have implemented hip surveillance programs [60]. The aim of this project is to establish such a monitoring program in Switzerland. Currently, the data set and the data collection procedure are being validated in Zurich. |
| Dr. M. Adams, PD Dr. A. Tscherter, Zurich, Bern | **Swiss Neonatal Network & Follow-up Group (SwissNeoNet):** Partial pseudonymized data from SwissNeoNet and Swiss-CP-Reg are regularly linked to identify missing children and facilitate collaborative research. In the first data linkage in August 2021, we could identify 186 preterm children not yet included in Swiss-CP-Reg. |
| Dr. A. Girardet, PD Dr. J. Fluss, Geneva | **Risk factors and outcomes of unilateral CP:** Unilateral CP is the most common form of CP affecting up to 40% of patients [55]. This project assesses different motor and cognitive patterns in Swiss children with unilateral CP. The results are compared with imaging, clinical, and socio-demographic data. |

We pursue participation in international research projects, in particular with the SCPE. CK, member of the Swiss-CP-Reg steering board, is a part of the SCPE. He founded a CP registry in the canton of St. Gallen in 2013. He recruited most of the children and adolescents with CP born between 1995 and 2011 and collected their data [2]. The Swiss-CP-Reg participates in projects from the SCPE with the data from St. Gallen [54].

## DISCUSSION

Swiss-CP-Reg led to standardised and harmonised collection of data on people with CP throughout Switzerland, facilitates data comparison between patients, regions, and countries, and has become a successful communication platform and a powerful tool for answering clinically relevant questions in CP. By August 2021, 546 patients have been enrolled.

### Comparison with other registries

The aims of Swiss-CP-Reg are comparable to those of other CP registries [55], but in addition include the creation of a research platform and a network for knowledge exchange. Swiss-CP-Reg uses the SCPE definition of CP like most registries (64%) [55], which facilitates comparison. Many (46%) registries require a minimum age of survival (44%) or a minimum severity criterion (46%) for inclusion [55]. We do not restrict on these criteria to ensure the inclusion of severely affected patients, who deceased at young age, and of mild cases. This reduces selection bias. We collect a complete data set after consent. Other registries use permission by government body or opt-off options [56], which makes recruitment easier. Most CP registries are governmentally funded (72%), others are supported by non-profit organizations/charities (12%) as Swiss-CP-Reg [55].

Preliminary results show that CP classification, severity, and prevalence of comorbidities are in line with other European countries [5,57-59]. Most participants are males, are diagnosed with a spastic bilateral CP subtype, and have mild motor impairment (GMFCS levels I or II) in line of expectation [5,57,58].

### Strengths and limitations

The successful start of the Swiss-CP-Reg is in large parts indebted to its steering board. It has been a bottom-up approach with all large Swiss clinics being represented. We strived to reach consensus on a fully standardised and complete data collection. The board did also set up a thorough methodology for patient identification and registration, individualised for each clinic. The continuous motivation of local medical staff and optimization of recruitment and data collection by steering board members is important since recruitment and data collection is time consuming. All steering board members are dedicated and have a broad national and international network, which facilitates funding and connections with all stakeholders. This, together with the dissemination activity of the registry, creates a platform for interdisciplinary and interregional communication and knowledge exchange, one of the key goals of the registry. Another strength of the Swiss-CP-Reg is its role as research platform. Several other factors contribute to its success. First, we developed our data set based on the data set established by SCPE, which facilitates comparison with other registries. Secondly, Swiss-CP-Reg simplifies research projects, as it obtained a broad ethical approval via a single informed consent to collect data from multiple sources, conduct surveys, and link data. We can also contact participants and their families directly to invite them for nested research. Finally, the registry can share partial coded data with research projects when they meet all legal requirements.

A limitation of the Swiss-CP-Reg is that it currently misses patients, who are diagnosed and treated in smaller clinics and private practices, and adult patients. This might have biased the distribution of severity, as it is possible that university hospitals treat more severe cases than smaller clinics and private practices. Efforts are made to become more inclusive by including all eligible patients. This will allow to make reliable statements about CP in Switzerland in the future and enable comparison with registries who apply different inclusion criteria, e.g., minimal severity of CP or exclusion of severely affected patients deceasing at young age. Secondly, the registry is limited now by retrospective data collection at time of recruitment. This will improve as soon as all patients already in follow-up have been included, so that recruitment can focus on incident cases, where data can be collected prospectively.

### Future developments

In the coming years patient recruitment will be expanded to include more clinics and private practices. We will also expand recruitment to adult participants. We plan in-depth analyses of topics causing patients with CP and their families concern. Data will be collected via questionnaires. The studied topics will depend on the outcome of our current project on needs and wishes of patients with CP and their families. The aim of these surveys will be to support CP patients and their families by uncovering possible intervention strategies for each topic. An important focus of research is the participation of people with CP in daily activities. Children and adolescents with CP participate less in social life than their peers [9]. Participation can be increased by therapeutic measures, but data on the situation in Switzerland is lacking. Therefore, we are currently setting up a study to investigate carriers and facilitators for the participation of children with CP. This study will assess how children with CP and their families evaluate their participation in social life. In addition, future research projects of Swiss-CP-Reg aim to understand the aetiology of CP; uncover prognostic factors of CP during the perinatal phase; evaluate the occurrence of associated syndromes and anomalies; determine comorbidities; evaluate and improve treatments and therapies; detect regional differences of medical coverage, evaluate the impact of socio-economic factors on diagnosis, prognosis, and quality of life; develop strategies to enhance quality of life; uncover needs and concerns of patients and families and evaluate early education and integration of CP patients in the school system and working environment. Further this data can be used for education of young medical staff members to train their diagnostic and therapeutic knowledge and skills to ensure improvement the care of patients suffering from cerebral palsy in future.

## CONCLUSIONS

The Swiss-CP-Reg is now established in Switzerland, generates rich information on people living with CP and has facilitated the development of national and international research projects in various fields. Swiss-CP-Reg serves as an instrument for medical care, clinical and epidemiological research, the assessment of health care structures, and the evaluation of medical procedures to increase our understanding of cerebral palsy, explore causes of CP and associated conditions, and improve clinical practice.

## Data Availability

All data produced in the present work are contained in the manuscript. As the cohort is ongoing, new or updated data is available upon reasonable request to the authors.

## ACKNOWLEDGMENTS

The authors express their gratitude to Dr. Christoph Kuenzle who initiated the Swiss-CP-Reg and all local hubs of Swiss Research Network of Clinical Paediatric Hubs (SwissPedNet) for their support for patient recruitment and data collection. We thank all patients in the Swiss-CP-Reg and their families for taking part. We thank all the physicians and local study teams who helped identify and register patients and worked closely with us throughout building the Swiss-CP-Reg. We are grateful to Prof. Dr. Thomas Dreher, Dr. Anne Girardet, Prof. Dr. Hubertus van Hedel, and Prof. Dr Christina Schulze for their collaboration on various projects and to the SACD for hosting the general assembly of Swiss-CP-Reg during their annual conference. We are thankful to our sponsors (see ‘Funding’), without whom this work would not be possible. SwissPedReg, member of the SwissPedNet, supports the Swiss-CP-Reg with its research infrastructure.

## ABBREVIATIONS

CFCS: Communication Function Classification System
CP: Cerebral Palsy
FSO: Federal Statistics Office
GMFCS: Gross Motor Function Classification System
ISPM: Institute of Social and Preventive Medicine
IQ: Intelligence quotient
MACS: Manual Ability Classification System
REDCap: Research Electronic Data Capture
SACD: Swiss Academy of Childhood Disability
SCPE: Surveillance of Cerebral Palsy in Europe
Swiss-CP-Reg: Swiss Cerebral Palsy Registry
SwissNeoNet: Swiss Neonatal Network and Follow-up Group
SwissPedNet: Swiss Research Network of Clinical Paediatric Hubs
SwissPedReg: Swiss Research Platform for Paediatric Registries

## REFERENCES

1. Graham, H.K.; Rosenbaum, P.; Paneth, N.; Dan, B.; Lin, J.P.; Damiano, D.L.; Becher, J.G.; Gaebler-Spira, D.; Colver, A.; Reddihough, D.S., et al. Cerebral palsy. Nature reviews. Disease primers 2016, 2, 15082.

2. Kuenzle, C.; Stojicevic, V.; Forni, R.; van Son, C.; Maier, O. Epidemiology of cerebral palsy in eastern switzerland (st. Gallen). Developmental Medicine & Child Neurology 2015, 57, 49–49.

3. Rosenbaum, P.; Paneth, N.; Leviton, A.; Goldstein, M.; Bax, M.; Damiano, D.; Dan, B.; Jacobsson, B. A report: The definition and classification of cerebral palsy april 2006. Developmental medicine and child neurology. Supplement 2007, 109, 8–14.

4. Hollung, S.J.; Bakken, I.J.; Vik, T.; Lydersen, S.; Wiik, R.; Aaberg, K.M.; Andersen, G.L. Comorbidities in cerebral palsy: A patient registry study. Developmental medicine and child neurology 2020, 62, 97–103.

5. Jonsson, U.; Eek, M.N.; Sunnerhagen, K.S.; Himmelmann, K. Cerebral palsy prevalence, subtypes, and associated impairments: A population-based comparison study of adults and children. Developmental medicine and child neurology 2019, 61, 1162–1167.

6. Fauconnier, J.; Dickinson, H.O.; Beckung, E.; Marcelli, M.; McManus, V.; Michelsen, S.I.; Parkes, J.; Parkinson, K.N.; Thyen, U.; Arnaud, C., et al. Participation in life situations of 8-12 year old children with cerebral palsy: Cross sectional european study. BMJ (Clinical research ed.) 2009, 338, b1458.

7. Michelsen, S.I.; Flachs, E.M.; Uldall, P.; Eriksen, E.L.; McManus, V.; Parkes, J.; Parkinson, K.N.; Thyen, U.; Arnaud, C.; Beckung, E., et al. Frequency of participation of 8-12-year-old children with cerebral palsy: A multi-centre cross-sectional european study. European journal of paediatric neurology : EJPN : official journal of the European Paediatric Neurology Society 2009, 13, 165–177.

8. Colver, A.; Thyen, U.; Arnaud, C.; Beckung, E.; Fauconnier, J.; Marcelli, M.; McManus, V.; Michelsen, S.I.; Parkes, J.; Parkinson, K., et al. Association between participation in life situations of children with cerebral palsy and their physical, social, and attitudinal environment: A cross-sectional multicenter european study. Archives of physical medicine and rehabilitation 2012, 93, 2154–2164.

9. Michelsen, S.I.; Flachs, E.M.; Damsgaard, M.T.; Parkes, J.; Parkinson, K.; Rapp, M.; Arnaud, C.; Nystrand, M.; Colver, A.; Fauconnier, J., et al. European study of frequency of participation of adolescents with and without cerebral palsy. European journal of paediatric neurology : EJPN : official journal of the European Paediatric Neurology Society 2014, 18, 282–294.

10. Colver, A.; Rapp, M.; Eisemann, N.; Ehlinger, V.; Thyen, U.; Dickinson, H.O.; Parkes, J.; Parkinson, K.; Nystrand, M.; Fauconnier, J., et al. Self-reported quality of life of adolescents with cerebral palsy: A cross-sectional and longitudinal analysis. Lancet (London, England) 2015, 385, 705–716.

11. Bundesamt für Sozialversicherungen BSV. Schweizerische eidgenossenschaft. Die medizinischen massnahmen in der invaliden-und krankenversicherung. https://www.parlament.ch/centers/documents/de/med-massnahmen-iv-kv-2013-03-15-d.pdf (14.07.2021),

12. Brehaut, J.C.; Kohen, D.E.; Raina, P.; Walter, S.D.; Russell, D.J.; Swinton, M.; O’Donnell, M.; Rosenbaum, P. The health of primary caregivers of children with cerebral palsy: How does it compare with that of other canadian caregiversã Pediatrics 2004, 114, e182–191.

13. Raina, P.; O’Donnell, M.; Rosenbaum, P.; Brehaut, J.; Walter, S.D.; Russell, D.; Swinton, M.; Zhu, B.; Wood, E. The health and well-being of caregivers of children with cerebral palsy. Pediatrics 2005, 115, e626–636.

14. Yamaoka, Y.; Tamiya, N.; Moriyama, Y.; Sandoval Garrido, F.A.; Sumazaki, R.; Noguchi, H. Mental health of parents as caregivers of children with disabilities: Based on japanese nationwide survey. PloS one 2015, 10, e0145200.

15. Lach, L.M.; Kohen, D.E.; Garner, R.E.; Brehaut, J.C.; Miller, A.R.; Klassen, A.F.; Rosenbaum, P.L. The health and psychosocial functioning of caregivers of children with neurodevelopmental disorders. Disabil Rehabil 2009, 31, 741–752.

16. Sipal, R.F.; Schuengel, C.; Voorman, J.M.; Van Eck, M.; Becher, J.G. Course of behaviour problems of children with cerebral palsy: The role of parental stress and support. Child Care Health Dev 2010, 36, 74–84.

17. Majnemer, A.; Shevell, M.; Law, M.; Poulin, C.; Rosenbaum, P. Indicators of distress in families of children with cerebral palsy. Disabil Rehabil 2012, 34, 1202–1207.

18. Park, M.S.; Chung, C.Y.; Lee, K.M.; Sung, K.H.; Choi, I.H.; Kim, T.W. Parenting stress in parents of children with cerebral palsy and its association with physical function. Journal of pediatric orthopedics. Part B 2012, 21, 452–456.

19. Tseng, M.H.; Chen, K.L.; Shieh, J.Y.; Lu, L.; Huang, C.Y.; Simeonsson, R.J. Child characteristics, caregiver characteristics, and environmental factors affecting the quality of life of caregivers of children with cerebral palsy. Disabil Rehabil 2016, 38, 2374–2382.

20. Woodgate, R.L.; Edwards, M.; Ripat, J.D.; Rempel, G.; Johnson, S.F. Siblings of children with complex care needs: Their perspectives and experiences of participating in everyday life. Child Care Health Dev 2016, 42, 504–512.

21. The cerebral palsy register of attica. https://eu-rd-platform.jrc.ec.europa.eu/scpe/scpe-network/members/greece_en (22.09.2021),

22. The cerebral palsy register of central italy - viterbo. https://eu-rd-platform.jrc.ec.europa.eu/scpe/scpe-network/members/italy_en (22.09.2021),

23. The cerebral palsy register of innsbruck. https://eu-rd-platform.jrc.ec.europa.eu/node/10_sl (22.09.2021),

24. Australian Cerebral Palsy Register, G. Australia and the australian cerebral palsy register for the birth cohort 1993 to 2006. Dev Med Child Neurol 2016, 58 Suppl 2, 3–4.

25. Shevell, M.I.; Dagenais, L.; Hall, N.; Consortium, R. Comorbidities in cerebral palsy and their relationship to neurologic subtype and gmfcs level. Neurology 2009, 72, 2090–2096.

26. Alriksson-Schmidt, A.; Rimstedt, A.B.; Hagglund, G. Cpup-a multidisciplinary secondary prevention program for individuals with cerebral palsy. Int J Integr Care 2019, 19.

27. Uldall, P.; Michelsen, S.I.; Topp, M.; Madsen, M. The danish cerebral palsy registry. A registry on a specific impairment. Dan Med Bull 2001, 48, 161–163.

28. Yim, S.Y.; Yang, C.Y.; Park, J.H.; Kim, M.Y.; Shin, Y.B.; Kang, E.Y.; Lee, Z.I.; Kwon, B.S.; Chang, J.C.; Kim, S.W., et al. Korean database of cerebral palsy: A report on characteristics of cerebral palsy in south korea. Ann Rehabil Med 2017, 41, 638–649.

29. Badawi, N.; Honan, I.; Finch-Edmondson, M.; Hogan, A.; Fitzgerald, J.; Imms, C. The australian & new zealand cerebral palsy strategy. Dev Med Child Neurol 2020, 62, 885.

30. Bufteac, E.G.; Andersen, G.L.; Torstein, V.; Jahnsen, R. Cerebral palsy in moldova: Subtypes, severity and associated impairments. Bmc Pediatr 2018, 18.

31. Almasri, N.A.; Saleh, M.; Abu-Dahab, S.; Malkawi, S.H.; Nordmark, E. Development of a cerebral palsy follow-up registry in jordan (cpup-jordan). Child Care Health Dev 2018, 44, 131–139.

32. Surveillance of Cerebral Palsy in Europe. Surveillance of cerebral palsy in europe: A collaboration of cerebral palsy surveys and registers. Surveillance of cerebral palsy in europe (scpe). Dev Med Child Neurol 2000, 42, 816–824.

33. Hurvitz, E.A.; Gross, P.H.; Gannotti, M.E.; Bailes, A.F.; Horn, S.D. Registry-based research in cerebral palsy: The cerebral palsy research network. Phys Med Rehabil Clin N Am 2020, 31, 185–194.

34. Wiedemann, A.; Pastore-Wapp, M.; Slavova, N.; Steiner, L.; Weisstanner, C.; Regényi, M.; Steinlin, M.; Grunt, S. Impact of stroke volume on motor outcome in neonatal arterial ischemic stroke. European journal of paediatric neurology : EJPN : official journal of the European Paediatric Neurology Society 2020, 25, 97–105.

35. Caspar-Teuscher, M.; Studer, M.; Regényi, M.; Steinlin, M.; Grunt, S. Health related quality of life and manual ability 5 years after neonatal ischemic stroke. European journal of paediatric neurology : EJPN : official journal of the European Paediatric Neurology Society 2019, 23, 716–722.

36. Grunt, S.; Mazenauer, L.; Buerki, S.E.; Boltshauser, E.; Mori, A.C.; Datta, A.N.; Fluss, J.; Mercati, D.; Keller, E.; Maier, O., et al. Incidence and outcomes of symptomatic neonatal arterial ischemic stroke. Pediatrics 2015, 135, e1220–1228.

37. Grunt, S.; Newman, C.J.; Saxer, S.; Steinlin, M.; Weisstanner, C.; Kaelin-Lang, A. The mirror illusion increases motor cortex excitability in children with and without hemiparesis. Neurorehabilitation and neural repair 2017, 31, 280–289.

38. Bruchez, R.; Jequier Gygax, M.; Roches, S.; Fluss, J.; Jacquier, D.; Ballabeni, P.; Grunt, S.; Newman, C.J. Mirror therapy in children with hemiparesis: A randomized observer-blinded trial. Developmental medicine and child neurology 2016, 58, 970–978.

39. Weisstanner, C.; Saxer, S.; Wiest, R.; Kaelin-Lang, A.; Newman, C.J.; Steinlin, M.; Grunt, S. The neuronal correlates of mirror illusion in children with spastic hemiparesis: A study with functional magnetic resonance imaging. Swiss medical weekly 2017, 147, w14415.

40. Datta, A.N.; Furrer, M.A.; Bernhardt, I.; Hüppi, P.S.; Borradori-Tolsa, C.; Bucher, H.U.; Latal, B.; Grunt, S.; Natalucci, G. Fidgety movements in infants born very preterm: Predictive value for cerebral palsy in a clinical multicentre setting. Developmental medicine and child neurology 2017, 59, 618–624.

41. van Hedel, H.J.A.; Severini, G.; Scarton, A.; O’Brien, A.; Reed, T.; Gaebler-Spira, D.; Egan, T.; Meyer-Heim, A.; Graser, J.; Chua, K., et al. Advanced robotic therapy integrated centers (artic): An international collaboration facilitating the application of rehabilitation technologies. Journal of neuroengineering and rehabilitation 2018, 15, 30.

42. Meyer-Heim, A.; Ammann-Reiffer, C.; Schmartz, A.; Schäfer, J.; Sennhauser, F.H.; Heinen, F.; Knecht, B.; Dabrowski, E.; Borggraefe, I. Improvement of walking abilities after robotic-assisted locomotion training in children with cerebral palsy. Archives of disease in childhood 2009, 94, 615–620.

43. Ammann-Reiffer, C.; Bastiaenen, C.H.; Meyer-Heim, A.D.; van Hedel, H.J. Effectiveness of robot-assisted gait training in children with cerebral palsy: A bicenter, pragmatic, randomized, cross-over trial (pelogait). BMC pediatrics 2017, 17, 64.

44. Rutz, E.; Vavken, P.; Camathias, C.; Haase, C.; Jünemann, S.; Brunner, R. Long-term results and outcome predictors in one-stage hip reconstruction in children with cerebral palsy. The Journal of bone and joint surgery. American volume 2015, 97, 500–506.

45. Brégou Bourgeois, A.; Mariani, B.; Aminian, K.; Zambelli, P.Y.; Newman, C.J. Spatio-temporal gait analysis in children with cerebral palsy using, foot-worn inertial sensors. Gait & posture 2014, 39, 436–442.

46. Bonnefoy-Mazure, A.; De Coulon, G.; Armand, S. Self-perceived gait quality in young adults with cerebral palsy. Developmental medicine and child neurology 2020, 62, 868–873.

47. Brunner, A.L.; Rutz, E.; Juenemann, S.; Brunner, R. Continuous vs. Blocks of physiotherapy for motor development in children with cerebral palsy and similar syndromes: A prospective randomized study. Developmental neurorehabilitation 2014, 17, 426–432.

48. Weber, P.; Bolli, P.; Heimgartner, N.; Merlo, P.; Zehnder, T.; Kätterer, C. Behavioral and emotional problems in children and adults with cerebral palsy. European journal of paediatric neurology : EJPN : official journal of the European Paediatric Neurology Society 2016, 20, 270–274.

49. Araneda, R.; Sizonenko, S.V.; Newman, C.J.; Dinomais, M.; Le Gal, G.; Ebner-Karestinos, D.; Paradis, J.; Klöcker, A.; Saussez, G.; Demas, J., et al. Protocol of changes induced by early hand-arm bimanual intensive therapy including lower extremities (e-habit-ile) in pre-school children with bilateral cerebral palsy: A multisite randomized controlled trial. BMC neurology 2020, 20, 243.

50. Aurich-Schuler, T.; Warken, B.; Graser, J.V.; Ulrich, T.; Borggraefe, I.; Heinen, F.; Meyer-Heim, A.; van Hedel, H.J.; Schroeder, A.S. Practical recommendations for robot-assisted treadmill therapy (lokomat) in children with cerebral palsy: Indications, goal setting, and clinical implementation within the who-icf framework. Neuropediatrics 2015, 46, 248–260.

51. Surveillance of Cerebral Palsy in Europe. Surveillance of cerebral palsy in europe (scpe): Scientific report 1998-2018; https://eu-rd-platform.jrc.ec.europa.eu/scpe_en, 2018.

52. Harris, P.A.; Taylor, R.; Minor, B.L.; Elliott, V.; Fernandez, M.; O’Neal, L.; McLeod, L.; Delacqua, G.; Delacqua, F.; Kirby, J., et al. The redcap consortium: Building an international community of software platform partners. Journal of biomedical informatics 2019, 95, 103208.

53. Harris, P.A.; Taylor, R.; Thielke, R.; Payne, J.; Gonzalez, N.; Conde, J.G. Research electronic data capture (redcap)--a metadata-driven methodology and workflow process for providing translational research informatics support. Journal of biomedical informatics 2009, 42, 377–381.

54. Sellier, E.; Goldsmith, S.; McIntyre, S.; Perra, O.; Rackauskaite, G.; Badawi, N.; Fares, A.; Smithers-Sheedy, H.; Europe, S.C.P.; Register, A.C.P. Cerebral palsy in twins and higher multiple births: A europe-australia population-based study. Developmental Medicine and Child Neurology 2021, 63, 712–720.

55. Ashwal, S.; Russman, B.S.; Blasco, P.A.; Miller, G.; Sandler, A.; Shevell, M.; Stevenson, R.; Quality Standards Subcommittee of the American Academy of, N.; Practice Committee of the Child Neurology, S. Practice parameter: Diagnostic assessment of the child with cerebral palsy: Report of the quality standards subcommittee of the american academy of neurology and the practice committee of the child neurology society. Neurology 2004, 62, 851–863.

56. Goldsmith, S.; McIntyre, S.; Smithers-Sheedy, H.; Blair, E.; Cans, C.; Watson, L.; Yeargin-Allsopp, M.; Australian Cerebral Palsy Register, G. An international survey of cerebral palsy registers and surveillance systems. Dev Med Child Neurol 2016, 58 Suppl 2, 11–17.

57. Surveillance of Cerebral Palsy in Europe (SCPE). Prevalence and characteristics of children with cerebral palsy in europe. Developmental medicine and child neurology 2002, 44, 633–640.

58. Andersen, G.L.; Irgens, L.M.; Haagaas, I.; Skranes, J.S.; Meberg, A.E.; Vik, T. Cerebral palsy in norway: Prevalence, subtypes and severity. European journal of paediatric neurology : EJPN : official journal of the European Paediatric Neurology Society 2008, 12, 4–13.

59. Oskoui, M.; Coutinho, F.; Dykeman, J.; Jette, N.; Pringsheim, T. An update on the prevalence of cerebral palsy: A systematic review and meta-analysis. Dev Med Child Neurol 2013, 55, 509–519.

60. Robb, J.E.; Hägglund, G. Hip surveillance and management of the displaced hip in cerebral palsy. Journal of children’s orthopaedics 2013, 7, 407–413.

